# Online Misinformation Susceptibility Scale: An adapted version for health-related misinformation

**DOI:** 10.1101/2025.09.11.25335589

**Authors:** A. Katsiroumpa, O. Konstantakopoulou, P. Gallos, I. Moisoglou, P. Mangoulia, O. Galani, M. Tsiachri, P. Galanis

**Author notes:** Corresponding author: P. Galanis. Funding: none. Conflicts of interest: none.

## Abstract

**OBJECTIVE:** To examine the validity and reliability of the adapted version of the Online Misinformation Susceptibility Scale (OMISS) for health-related misinformation.

**METHOD:** We examined the reliability of the Health-Related Online Misinformation Susceptibility Scale (HR-OMISS) by calculating Cronbach alpha and McDonald Omega. We examined the construct validity of the HR-OMISS by performing confirmatory factor analysis. We examined the concurrent validity of the HR-OMISS using the Trust in Scientists Scale, the single-item scientists confidence scale, the Conspiracy Mentality Questionnaire (CMQ), and the single item conspiracy belief. We examined known-groups validity of the HR-OMISS by comparing healthcare workers with a MSc diploma versus those without one.

**RESULTS:** We found that the HR-OMISS had very good reliability since Cronbach coefficient alpha 0.920 and McDonald Omega was 0.922. We found that the adapted version of the OMISS for health-related misinformation (HR-OMISS) had a one-factor structure as the original version (OMISS). Concurrent validity of the HR-OMISS was very good since we found statistically significant negative correlation between the HR-OMISS and the Trust in Scientists Scale (r = -0.330, p-value < 0.001), and the single-item scientists confidence scale (r = -0.258, p-value < 0.001). Moreover, we found a positive correlation between the HR-OMISS and the CMQ (r = 0.118, p-value = 0.072) and the single item conspiracy belief (r = 0.074, p-value = 0.260). The HR-OMISS showed known-groups validity since the mean score for healthcare workers with a MSc diploma (25.6) was lower than those without a MSc diploma (29.6), (p-value = 0.002).

**CONCLUSIONS:** The HR-OMISS is a valid and reliable tool to measure levels of health-related online misinformation susceptibility.

## Introduction

Verifying information and identifying misinformation are essential skills for enhancing our understanding and making informed decisions, as misinformation can distort perceptions and generate confusion about reality.^1,2^ Misinformation refers to any information that is clearly false or misleading, irrespective of its origin or intent. It is important to distinguish misinformation from disinformation, the latter being false information that is intentionally created and disseminated to deceive.^3,4^

Misinformation is particularly concerning because it promotes false beliefs and can exacerbate partisan divisions, even concerning basic facts. In online environments, individuals frequently encounter content presented as accurate, despite lacking verification or being outright incorrect. Exposure to such false narratives—even passively—can increase the likelihood that individuals will later accept them as true.^5^ The consequences of misinformation include a reduction in the accuracy of both general knowledge and memory of specific events.^6^ When people hold incorrect beliefs, they may express their opinions with unwarranted confidence, potentially shaping public discourse and influencing policy decisions. This type of misinformation is especially pervasive when it originates from sources perceived as “credible and authoritative,” despite the inaccuracy of the information being shared.^7^

Particular attention must be directed toward health-related misinformation found online, as it can significantly impact rational decision-making, influence health behaviors, and affect outcomes such as vaccine acceptance, timely access to care, and adherence to treatment.^8–10^ A recent umbrella review, encompassing 31 systematic reviews, revealed that the prevalence of health-related misinformation on social media varies widely—from 0.2% to 28.8%.^10^ This review highlighted that one of the most detrimental consequences of such misinformation is the increased likelihood of misinterpreting or misunderstanding established scientific evidence. Additionally, it can adversely affect mental health, lead to inefficient utilization of healthcare resources, and contribute to the rising trend of vaccine hesitancy.

The dissemination of unreliable health information can result in delayed medical intervention and contribute to divisive or harmful public discourse. A recent and striking example of the tangible public health consequences of misinformation is the COVID-19 pandemic. Studies from multiple countries have demonstrated a strong correlation between belief in COVID-19 misinformation and reduced compliance with public health measures, lower vaccine uptake, and decreased likelihood of recommending vaccination to others.^11–14^ Experimental findings indicate that exposure to vaccine-related misinformation led to an approximately six-percentage-point decline in vaccination intent among individuals who had initially expressed a firm intention to receive the vaccine, thereby jeopardizing efforts to achieve herd immunity.^15^ Furthermore, analyses of social media networks suggest that, without targeted intervention, anti-vaccine narratives could come to dominate online discussions over the next decade.^16^ Additional research has linked exposure to COVID-19 misinformation with the use of harmful substances^17^ and an increased risk of violent behavior.^18^ Notably, the public health impact of health-related misinformation predates the COVID-19 crisis. For example, the debunked claim connecting the MMR vaccine to autism significantly reduced vaccination rates in the United Kingdom.^19^

In this context, our aim was to examine the validity and reliability of the adapted version of the Online Misinformation Susceptibility Scale (OMISS)^20^ for health-related misinformation, namely the Health-Related Online Misinformation Susceptibility Scale (HR-OMISS). Moreover, we examined an optimal cut-off point for the HR-OMISS.

## Methods

### Study design

As developers of the OMISS suggest, we slightly adapted the tool to measure specifically health-related online misinformation susceptibility and not misinformation susceptibility in general.^20^ In particular, the only transformation we made on the OMISS is to slightly adapt the introductory note by adding two simple words, i.e. “Please think about what you do when you see a health-related post or story that interests you on social media or websites”. The nine items and answers of the OMISS remained the same.^20^ Thus, we examined the validity and reliability of an adapted version of the OMISS for health-related misinformation, i.e. the Health-Related Online Misinformation Susceptibility Scale (HR-OMISS).

We conducted a cross-sectional study in Greece with a sample of healthcare workers. We approached healthcare workers through Facebook groups. We developed an online version of the study questionnaire by using Google forms. Our sample included physicians, nurses, psychologists, pharmacists, midwifes, etc. Then, we invited healthcare workers participate in our study through an anonymous way since we did not collect personal data. An introduction note informed healthcare workers about the aim and the design of the study. Healthcare workers that agreed to participate in our study may then fill out the online questionnaire. We collected our data during October 2025.

We examined the reliability of the HR-OMISS by calculating Cronbach’s alpha and McDonald’s Omega. Cronbach’s alpha and McDonald’s Omega higher than 0.6 indicate acceptable internal reliability. Also, we performed a test-retest study with 62 healthcare workers to examine the reliability of the HR-OMISS by calculating the intraclass correlation coefficient.

We examined the construct validity of the HR-OMISS by performing confirmatory factor analysis.^21^ We examined the concurrent validity of the HR-OMISS using the Trust in Scientists Scale, the single-item scientists confidence scale, the Conspiracy Mentality Questionnaire (CMQ), and the single item conspiracy belief. We expected negative correlations between the HR-OMISS and the Trust in Scientists Scale, and the single-item scientists confidence scale. Moreover, we expected positive correlations between the HR-OMISS and the CMQ, and the single item conspiracy belief.

The HR-OMISS includes nine items and measures health-related online misinformation susceptibility. The introductory note in the HR-OMISS is the following: “Please think about what you do when you see a health-related post or story that interests you on social media or websites”. Example items are the followings: “How often do you check the website domain and URL?”, “How often do you check the publication date of the post?”. Answers are on a five-point Likert scale: never (5), rarely (4), sometimes (3), very often (2), always (1). Total score ranges from 9 to 45 by adding answers in nine items. Higher scores indicate higher health-related misinformation susceptibility.^20^

The Trust in Scientists Scale^22^ measure levels of trust in scientists. We used the valid Greek version of the Trust in Scientists Scale.^22^ We used one of the four factors of the scale, i.e. the integrity factor. The “integrity factor” comprises three items such as “How honest or dishonest are most scientists?”. Answers are on a five-point Likert scale from 1 (very dishonest) to 5 (very honest). Total integrity score ranges from 1 to 5. Higher values indicate higher levels of trust in scientists. In our study, Cronbach’s alpha for the Trust in Scientists Scale was 0.913, and McDonald’s Omega was 0.918.

The single-item scientists confidence scale is taken from the Pew Research Center, and measures level of confidence in scientists.^23^ The single-item scientists confidence scale includes the following question: “How much confidence do you have in the scientists to act in the best interests of the public?”. Answers are on a five-point Likert scale from 1 (no confidence at all) to 5 (a great deal of confidence). Higher scores indicate higher levels of trust in sciences.

The Conspiracy Mentality Questionnaire measures generic conspiracy beliefs and includes five items.^24^ Answers are on an 11-point Likert scale from 0 (totally disagree) to 10 (totally agree). Total score on the CMQ ranges from 0 to 50 with higher values indicate higher conspiracy beliefs. We used the valid Greek version of the CMQ.^25^ In our study, Cronbach’s alpha for the CMQ was 0.840, and McDonald’s Omega was 0.862.

We used the Trust in Scientists Scale and the Conspiracy Mentality Questionnaire as external criterions for the Receiver Operating Characteristic (ROC) analysis.

### Ethical considerations

We applied the guidelines of the Declaration of Helsinki to perform this study.^26^ Additionally, the study protocol was approved by the Ethics Committee of Faculty of Nursing, National and Kapodistrian University of Athens (approval number; 55, June 23, 2025).

### Statistical analysis

We performed confirmatory factor analysis (CFA) to examine the construct validity of the HR-OMISS. In particular, we calculated chi-square/degree of freedom (x^2^/df); root mean square error of approximation (RMSEA); goodness of fit index (GFI); adjusted goodness of fit index (AGFI); Tucker–Lewis index (TLI); incremental fit index (IFI); normed fit index (NFI); comparative fit index (CFI).^27,28^ Acceptable value for x^2^/df is <5, for RMSEA is <0.10, and for all other measures in the CFA >0.90. We used the AMOS version 21 (Amos Development Corporation, 2018) to conduct the CFA.

We calculated Pearson’s correlation coefficient between the HR-OMISS and the Trust in Scientists Scale, the single-item scientists confidence scale, the CMQ, and the single item conspiracy belief to examine the concurrent validity of the HR-OMISS. Also, we calculated intraclass correlation coefficients between the two HR-OMISS measurements in test-retest study.

We examined known-groups validity of the HR-OMISS by comparing healthcare workers with a MSc diploma versus those without one. We expected less misinformation susceptibility among those with a MSc diploma. We used the independent samples t-test to explore difference on the HR-OMISS score between the two groups.

We employed the Receiver Operating Characteristic analysis to identify an optimal cut-off point for the HR-OMISS using the Trust in Scientists Scale, and the Conspiracy Mentality Questionnaire as external criterions. We calculated sensitivity, specificity, and the Youden index. These measures take values from 0 to 1 with higher values indicating better diagnostic value of the HR-OMISS. The Youden index defines an optimal cut-off point and is calculated as (Sensitivity + Specificity) – 1. Additionally, we calculated the area under the curve (AUC), 95% confidence interval (CI), and p-value. Values for the AUC between 0.5 and 0.7 indicate low accuracy, values between 0.71 and 0.9 indicate moderate accuracy, and values greater than 0.9 indicate high accuracy.^29–31^ After defining the best cut-off point for the HR-OMISS, healthcare workers with a score above this value are considered to experience high levels of online health-related misinformation susceptibility.

P-values less than 0.05 were considered as statistically significant. We used the IBM SPSS 28.0 (IBM Corp. Released 2021. IBM SPSS Statistics for Windows, Version 28.0. Armonk, NY: IBM Corp) for the analysis.

## Results

Study population included 232 healthcare workers. Among them, 84.1% (n=195) were females and 15.9% (n=37) were males. Mean age of our sample was 38.3 years (standard deviation; 8.4), with a median of 38.5 years, a minimum age of 24 years, and a maximum age of 53 years.

We found that the HR-OMISS had very good reliability since Cronbach’s coefficient alpha 0.920 and McDonald’s Omega was 0.922. Moreover, intraclass correlation coefficient for the HR-OMISS in the test-retest study was 0.986 (95% confidence interval = 0.977 to 0.992, p-value < 0.001).

We found that the adapted version of the OMISS for health-related misinformation (HR-OMISS) had a one-factor structure as the original version (OMISS). All indices indicated an acceptable one-factor model (Figure 1). In particular, x^2^/df was 1.624, RMSEA was 0.052, GFI was 0.971, AGFI was 0.932, TLI was 0.983, IFI was 0.991, NFI was 0.977, and CFI was 0.991 (Table 1). Moreover, standardized regression weights for the six items ranged from 0.64 to 0.87.

Concurrent validity of the HR-OMISS was very good since we found statistically significant negative correlation between the HR-OMISS and the Trust in Scientists Scale (r = -0.330, p-value < 0.001), and the single-item scientists confidence scale (r = -0.258, p-value < 0.001). In other words, less confidence in scientists was related with more health-related online misinformation susceptibility. Moreover, we found a positive correlation between the HR-OMISS and the CMQ (r = 0.118, p-value = 0.072) and the single item conspiracy belief (r = 0.074, p-value = 0.260). In other words, healthcare workers who believed in conspiracy theories are those with higher levels of health-related online misinformation susceptibility.

The HR-OMISS showed known-groups validity since the mean score for healthcare workers with a MSc diploma (25.6, SD; 8.4) was lower than those without a MSc diploma (29.6, SD; 8.6), (p-value = 0.002). Thus, healthcare workers with a higher educational level were less susceptible to misinformation.

**Table 1.** Confirmatory factor analysis for the adapted version of the Online Misinformation Susceptibility Scale for health-related misinformation (HR-OMISS).

| Model | $\chi^2$ | df | $\chi^2/\text{df}$ | RMSEA | GFI | AGFI | TLI | IFI | NFI | CFI |
| --- | --- | --- | --- | --- | --- | --- | --- | --- | --- | --- |
| Nine items | 30.861 | 19 | 1.624 | 0.052 | 0.971 | 0.932 | 0.983 | 0.991 | 0.977 | 0.991 |

**Figure 1.**
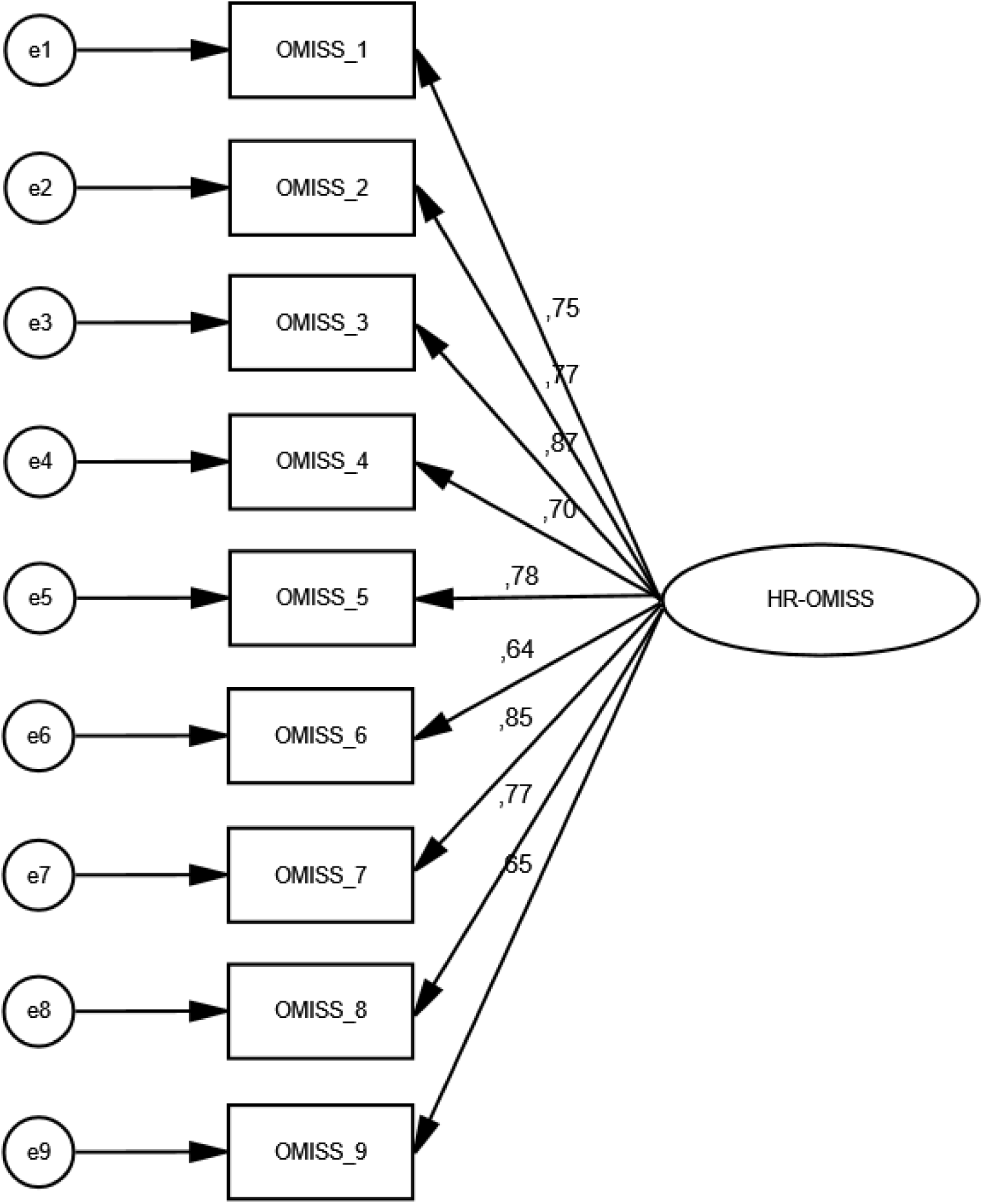
Confirmatory factor analysis for the adapted version of the Online Misinformation Susceptibility Scale for health-related misinformation (HR-OMISS).

We employed ROC analysis to define an optimal cut-off point for the HR-OMISS. We found that the best cut-off point for the HR-OMISS was 23 using the Trust in Scientists Scale, and the Conspiracy Mentality Questionnaire as external criterions (Table 2). Therefore, all external criterions suggested the same cut-off point (23) for the HR-OMISS. Youden’s index for the Trust in Scientists Scale, and the Conspiracy Mentality Questionnaire was 0.309 and 0.202, respectively. The AUC for the Trust in Scientists Scale (Figure 2), and the Conspiracy Mentality Questionnaire (Figure 3) was 0.696 and 0.610, respectively. Sensitivity for the Trust in Scientists Scale, and the Conspiracy Mentality Questionnaire was 0.736, and 0.676, respectively. Specificity for the Trust in Scientists Scale, and the Conspiracy Mentality Questionnaire was 0.573, and 0.525, respectively.

**Table 2.**
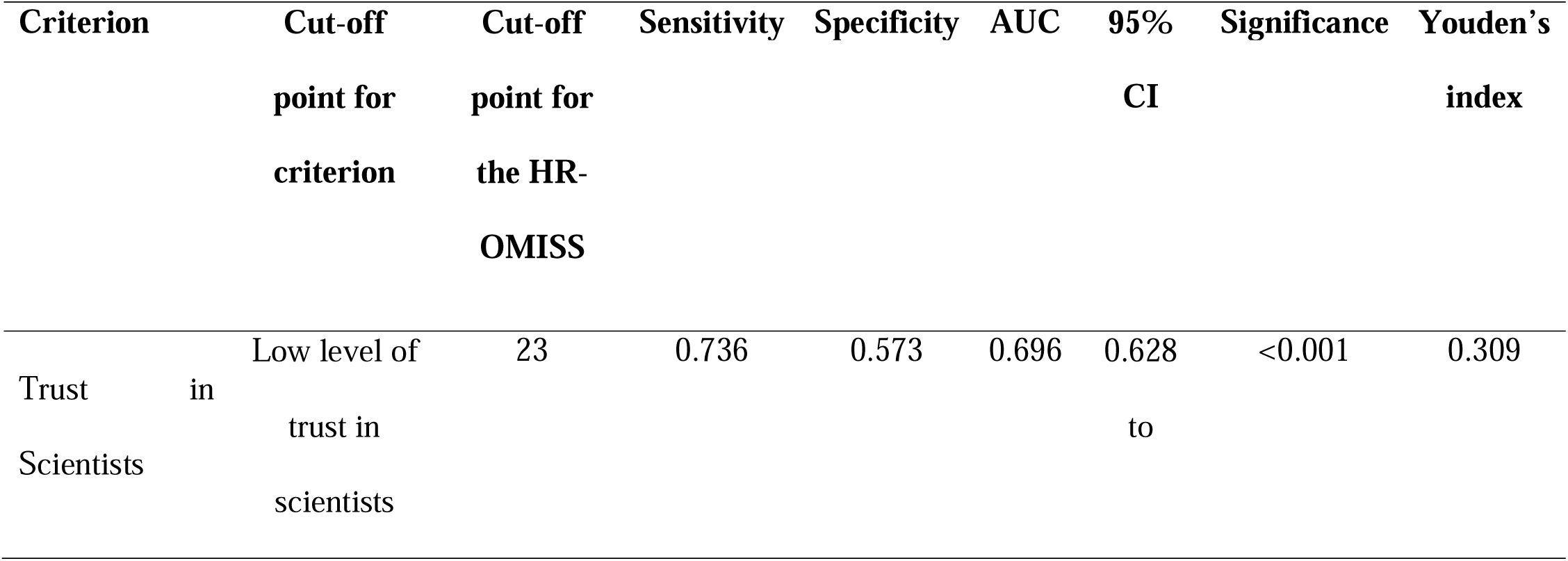
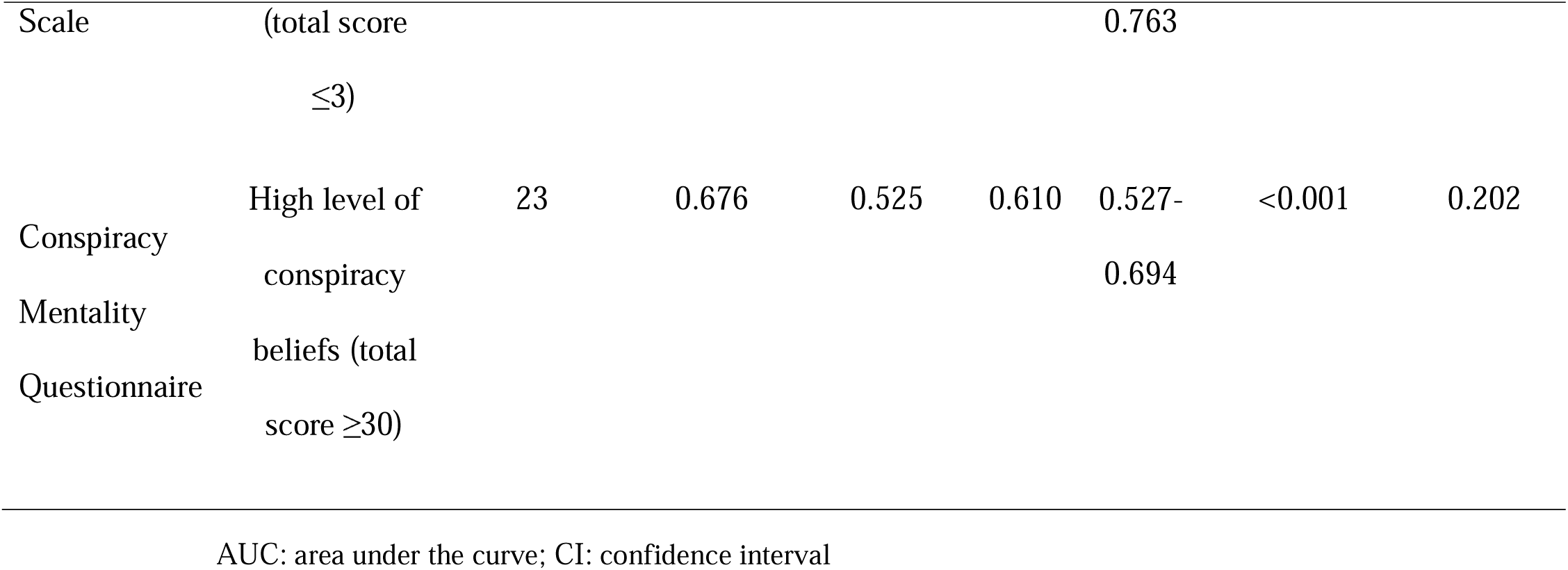
Predictive validity of the Health-Related Online Misinformation Susceptibility Scale (HR-OMISS).

**Figure 2.**
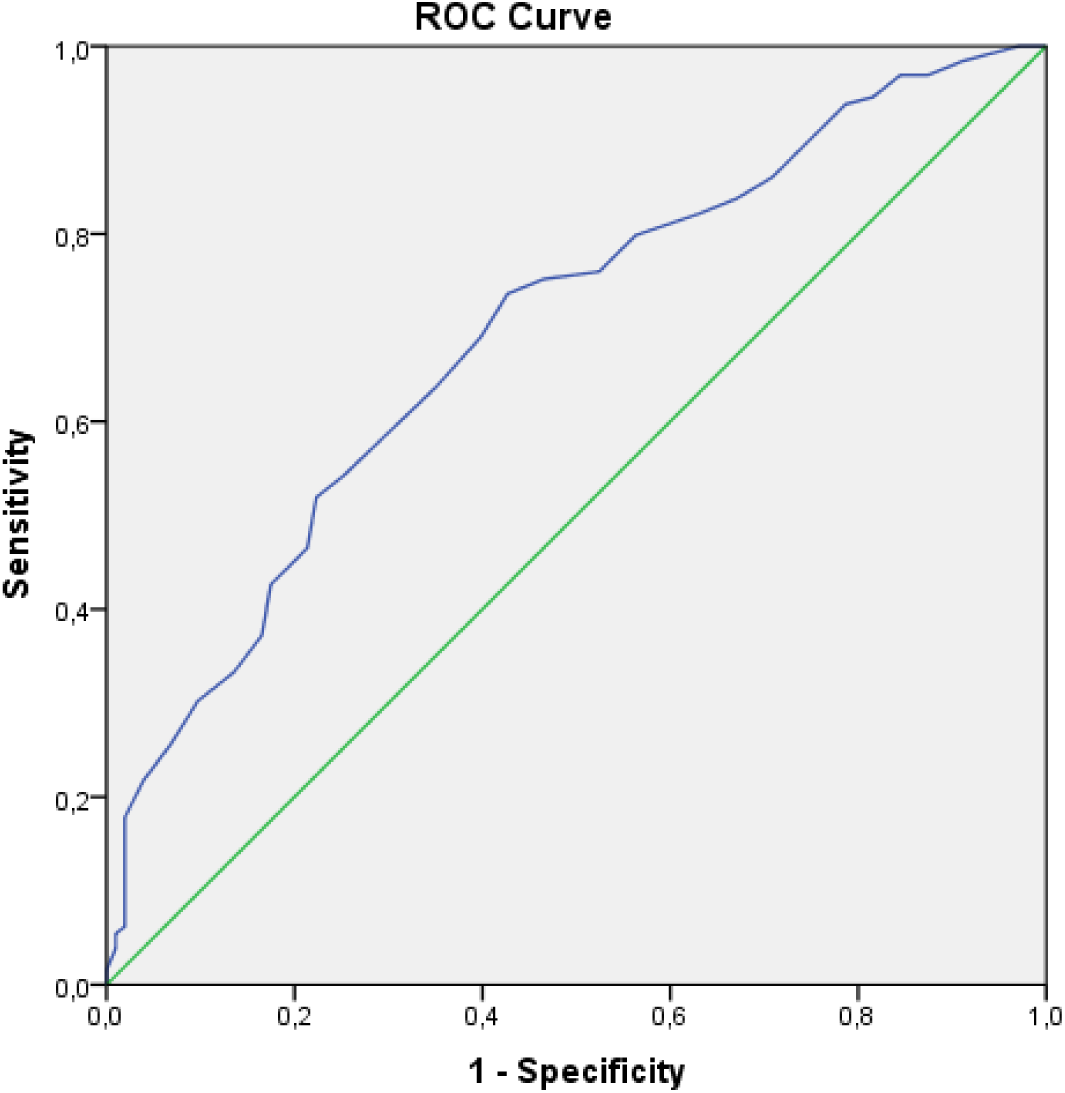
ROC curve of the Health-Related Online Misinformation Susceptibility Scale by using the Trust in Scientists Scale as the gold standard.

**Figure 3.**
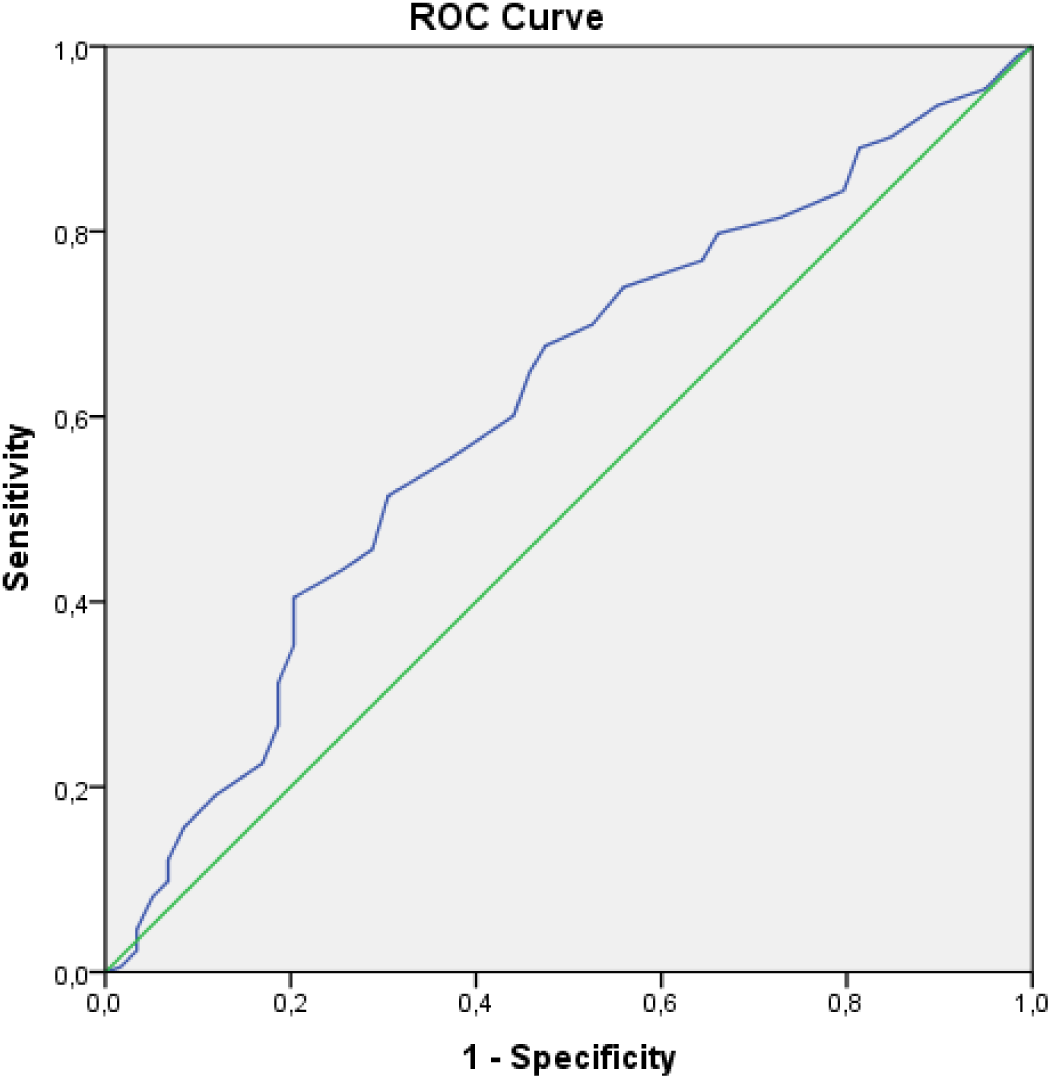
ROC curve of the Health-Related Online Misinformation Susceptibility Scale by using the Conspiracy Mentality Questionnaire as the gold standard.

## Discussion

Our findings showed that the HR-OMISS is a valid and reliable tool to measure levels of health-related online misinformation susceptibility.

We found that the HR-OMISS had very good psychometric properties. In particular, our confirmatory factor analysis confirmed the one-factor nine items structure of the original version (OMISS). Also, the HR-OMISS had very good concurrent validity since we found a negative correlation between the HR-OMISS and the Trust in Scientists Scale, and the single-item scientists confidence scale. Additionally, we found a positive correlation between the HR-OMISS and the CMQ and the single item conspiracy belief. Similarly, the HR-OMISS had very good reliability since Cronbach’s coefficient alpha and McDonald’s Omega were higher than 0.922. Moreover, intraclass correlation coefficient for the HR-OMISS in the test-retest study was 0.986.

Measurement of health-related misinformation susceptibility with valid scales such as the HR-OMISS could offer several important scientific, social, and practical benefits. Using validated tools like the HR-OMISS to measure susceptibility to health-related misinformation can provide significant scientific, societal, and practical advantages. Such measurement can enable healthcare professionals, institutions, and policymakers to respond more effectively to the growing challenge of misinformation in the digital age. Specifically, the HR-OMISS can enhance our capacity to detect individuals who are more prone to believing false health information. By identifying these vulnerable groups, policymakers can design and implement targeted, evidence-based public health campaigns—such as those addressing vaccine safety and efficacy—tailored to specific personality traits, cognitive styles, or emotional sensitivities.^32,33^ An example of this approach is the World Health Organization’s use of susceptibility assessments to guide its infodemic management efforts.^34^

Additionally, governments, public health authorities, and media outlets can tailor their communication strategies to reduce misinterpretation. For example, targeted media literacy initiatives led by reputable bodies, such as the European Commission, have demonstrated success in decreasing health misinformation on topics like the COVID-19 pandemic and vaccine hesitancy.^35^ Researchers can also employ health-related misinformation tools to evaluate the effectiveness of educational programs and interventions in diminishing belief in false information. Moreover, such tools can facilitate interdisciplinary research across fields such as healthcare, communication, political science, and public health, by providing a standardized and validated measure of susceptibility to misinformation.

This study has a few limitations that should be acknowledged. First, the use of a convenience sample from the population of healthcare workers for the validation of the HR-OMISS restricts the generalizability of the results. Future research should aim to validate the tool using more diverse and representative samples. Second, data were collected through self-reported questionnaires to assess the concurrent validity of the HR-OMISS. Further research is recommended to investigate additional forms of validity for the HR-OMISS. Finally, we examined the validity and reliability in a language (Greek). Thus, further studies should examine the psychometric properties of the scale in other languages and cultural settings.

In conclusion, the Greek version of the Health-Related Online Misinformation Susceptibility Scale demonstrated very good psychometric properties, making it a valid and reliable instrument for assessing levels of health-related online misinformation susceptibility.

## Data Availability

All data produced in the present study are available upon reasonable request to the authors

